# Evaluation of Some Serum Minerals in Type 2 Diabetic Patients Attending Federal Medical Centre Owo

**DOI:** 10.1101/2024.10.07.24315051

**Authors:** Fiyinfoluwa Olamide Ajao, Timothy God-Giveth Olusegun, Blessing Oluwatosinmile Oyeromi, Emmanuel Ifeanyi Obeagu

## Abstract

Minerals and trace elements play crucial roles in physiological processes, and alterations in their levels can have significant consequences for metabolic health. This study aimed to investigate serum mineral levels in patients with type 2 diabetes mellitus (T2DM) compared to healthy controls and explore their potential implications for disease pathophysiology. A total of 50 T2DM patients and 30 healthy individuals were included in the study. Serum levels of iron, copper, calcium, magnesium, and zinc were analysed using flame atomic absorption spectrometry. The results revealed significantly higher levels of iron and copper in T2DM patients compared to controls, while lower levels of calcium, magnesium, and zinc were observed in the T2DM group. These findings are consistent with previous research, highlighting the importance of mineral homeostasis in T2DM. Further analysis showed weak interrelationships among the studied minerals in T2DM patients, suggesting complex mechanisms underlying mineral metabolism in the disease. The discussion delved into potential mechanisms contributing to these alterations, including glycosuria-induced urinary loss of zinc and calciuresis due to hyperglycemia. Additionally, the study emphasised the importance of zinc and manganese in insulin production and release, as well as the contradictory findings regarding calcium levels in T2DM. Overall, this study provides valuable insights into serum mineral levels in T2DM and highlights the need for further research to elucidate their roles in disease progression and management.

## Introduction

Minerals and trace elements are micronutrients that have important biochemical functions primarily in metabolic processes, cell protection, gene regulation, hormone perception, signal transduction, and reproduction. Due to their role in maintaining cellular metabolic processes, their deficiencies disrupt normal physiological function, associating them with several health issues in humans. Alterations in mineral metabolism are observed in diabetes, suggesting a relationship between mineral homeostasis and disease pathophysiology. Thus, nutrients may have distinct functions in the development and advancement of this condition [1].

Type 2 diabetes mellitus (T2DM) represents a significant global health challenge characterised by inadequate insulin production or utilisation, resulting in chronic hyperglycemia and systemic complications [2]. It has resulted in a direct cause of 1.5 million deaths, with 48% of those deaths occurring before the age of 70. Additionally, raised blood glucose caused around 20% of cardiovascular deaths and 460,000 kidney disease deaths. These have been a burden, with a 3% increase in age-standardised mortality rates between 2000 and 2019 and a 13% increase in lower-middle-income countries [3].

Despite considerable advancements in diabetes management, the interplay between mineral homeostasis and T2DM pathophysiology remains under investigation. The study of essential minerals such as zinc, magnesium, calcium, copper, and iron reveals their potential roles in glucose homeostasis and diabetic complications. Specifically, zinc is crucial for insulin synthesis and secretion. A study conducted showed that administering zinc to diabetic rats resulted in an increase in the activity of the glutathione peroxidase enzyme and a decrease in the levels of malondialdehyde and nitric oxide, which validated the antioxidant properties of zinc in type 2 diabetes mellitus [4]. Magnesium deficits are frequently observed in poorly controlled T2DM patients.

Also, copper, an essential trace mineral, serves as a crucial cofactor required for redox reactions. An imbalance in its level can result in detrimental health consequences, as its oxidation varies and depends on the level of consumption. Therefore, it can serve as both an antioxidant and a pro-oxidant. The antioxidant properties of copper are evidenced by its role as a vital constituent of copper/zinc superoxide dismutase (SOD). Superoxide dismutase (SOD) functions by eliminating free radicals in every cell of the body, thereby providing protection against oxidative stress. Nevertheless, excessive copper can lead to insulin resistance by functioning as a pro-oxidant. An excess amount of copper stimulates the production of reactive oxygen species (ROS), resulting in heightened oxidative stress and ultimately contributing to the development of diabetes [5]. Calcium, traditionally recognised for its skeletal benefits, has emerged as a potential modulator of T2DM risk, with evidence suggesting a protective effect against its development [6].

Iron, a vital micronutrient, plays a crucial role in controlling the development and proliferation of cells and facilitates the transfer of electrons between cells, thereby influencing genomic synthesis. Blood glucose levels are intricately linked to the iron concentrations in islet beta cells. Due to the increased levels of iron transport proteins in the islet b-cells compared to other tissue cells, it is more probable for the islet b-cells to accumulate iron. An excess amount of iron can lead to an overabundance of oxidative stress, which can cause the death of islet b-cells and ultimately disrupt insulin secretion, increasing the likelihood of insulin resistance [7].

Despite the growing recognition of mineral imbalances in T2DM, the precise mechanisms and their clinical implications remain incompletely understood, particularly in African populations where T2DM prevalence is escalating. Addressing this knowledge gap is essential for devising targeted prevention and treatment strategies. This study aims to investigate; what are the serum levels of essential minerals (zinc, magnesium, calcium, copper, and iron) in patients with type 2 diabetes mellitus attending Federal Medical Centre Owo? Are there significant differences in serum mineral levels between diabetic patients with varying glycemic control and disease duration? Do alterations in serum mineral levels correlate with the presence of microvascular and macrovascular complications in patients with type 2 diabetes mellitus? By elucidating the relationship between serum mineral levels and T2DM characteristics, this research endeavours to contribute to a deeper understanding of mineral metabolism in diabetes and inform more targeted therapeutic interventions.

## Material and Methods

### Data collection

The study was conducted with the requisite ethical clearance obtained from the Federal Medical Centre, Owo, Nigeria, and comprised patients diagnosed with type 2 diabetes, with both male and female individuals included. The study sample consisted of patients with type 2 diabetes attending the Federal Medical Centre, Owo, Nigeria, including both male and female individuals, with 50 samples collected from type 2 diabetic patients and 30 non-diabetic healthy individuals included as control subjects. The inclusion criteria were informed consent and diagnosed type 2 diabetic patients for the case group, and informed consent and healthy subjects without type 2 diabetes mellitus for the control group, while non-diagnosed patients with type 2 diabetes and those with serious medical disorders were excluded. Five millilitres (5 ml) of venous blood were obtained from each subject by applying a tourniquet around the arm above the elbow, with the antecubital fossa disinfected with a 70% alcohol-soaked swab, and the blood was obtained aseptically and dispensed into lithium heparin anticoagulant bottles and plain bottles, then spun at 1200 rpm for 5 minutes to obtain plasma, which was stored in plain bottles at -20°C until analysed.

### Analytical Methods

The contents of Fe, Zn, and Cu in the serum were estimated using flame atomic absorption spectrometry (AAS-3 spectrometer with BC, Carl-Zeiss, Jena, Germany). The analytical procedure involved digesting serum samples using the conventional wet acid method, as described by Memon et al. (2007) [8]. Specifically, a whole serum sample was taken into a conical flask, followed by the addition of 3ml of a freshly prepared mixture of concentrated nitric acid and hydrogen peroxide (HNO3 + H2O2) (2:1 V/V). The mixture was left to stand for 10 minutes, then digested at 60-70°C for 1-2 hours. The digest was treated with 2ml nitric acid and a few drops of H2O2, while heating continued on a hot plate at about 80°C until a clear solution was obtained. The excess acid mixture was evaporated to a semi-dry mass, cooled, and diluted with distilled water to the mark of a volumetric flask (100ml). The resulting solution was transferred into a sample bottle. A blank extraction (without the sample) was carried out using triply distilled water, and the AAS machine (Buck Scientific 210 VGP) was powered for 45 minutes to 1 hour before analysis to increase its efficiency.

## Results

Table 3: Study subjects with Type 2 Diabetes Mellitus (T2DM) exhibited significantly higher levels of Iron (p < 0.001) and Copper (p = 0.001) compared to healthy controls. Conversely, lower levels of Calcium, Magnesium, and Zinc were observed in T2DM patients compared to controls (p < 0.05)

Table 2: The Pearson bivariate correlation analysis of serum minerals and trace elements in Type 2 diabetic (T2DM) subjects revealed weak interrelationships. Calcium (Ca) exhibited a weak negative correlation with copper (Cu) (r = -0.2918, p = 0.067707) and weak positive correlations with iron (Fe) (r = 0.061047, p = 0.708255), Manganese (Mn) (r = 0.226198, p = 0.160467), and Zinc (Zn) (r = 0.064467, p = 0.692689). Copper (Cu) showed a weak positive correlation with Iron (Fe) (r = 0.184958, p = 0.253215) and a weak negative correlation with Zinc (Zn) (r = -0.27933, p = 0.080885). Iron (Fe) displayed a weak positive correlation with Manganese (Mn) (r = 0.210632, p = 0.192027), while Manganese (Mn) showed a weak positive correlation with Zinc (Zn) (r = 0.157212, p = 0.212094). Overall, the correlations were mostly weak and not statistically significant, indicating limited associations among the studied minerals and trace elements in T2DM subjects.

**TABLE 1:** Comparison of Serum Mineral Levels between Control and Type 2 Diabetic Groups.

| Parameters | Control Subjects<br>N = 30 | Type 2 Diabetic Subjects<br>N= 50 | P-Value |
| --- | --- | --- | --- |
| Calcium<br>(mg/L) | 15.51 ± 1.69 | 11.35 ± 1.82 | 0.001 |
| Copper<br>(mg/L) | 0.15 ± 0.02 | 0.26 ± 0.08 | 0.001 |
| Iron<br>(mg/L) | 0.08 ± 0.02 | 0.16 ± 0.37 | 0.001 |
| Magnesium<br>(mg/L) | 0.10 ± 0.01 | 0.06 ± 0.07 | 0.003 |
| Zinc<br>(mg/L) | 0.99 ± 0.22 | 0.83 ± 0.20 | 0.001 |

Values were expressed as mean ± standard deviation. n=number of subjects. Significance level at p<0.05

**Table 2:** Pearson bivariate correlation analysis on serum Minerals and trace elements among diabetes study subjects.

|  |  | **Ca** | **Cu** | **Fe** | **Mn** | **Zn** |
| --- | --- | --- | --- | --- | --- | --- |
| **Ca** | **r** | 1 | -0.2918 | 0.061047 | 0.226198 | 0.064467 |
|  | *p*-Value |  | 0.067707 | 0.708255 | 0.160467 | 0.692689 |
| **Cu** | **r** | -0.2918 | 1 | 0.184958 | -0.01057 | -0.27933 |
|  | *p*-Value | 0.067707 |  | 0.253215 | 0.948377 | 0.080885 |
| **Fe** | **r** | 0.061047 | 0.184958 | 1 | 0.210632 | 0.157212 |
|  | *p*-Value | 0.708255 | 0.253215 |  | 0.192027 | 0.332639 |
| **Mn** | **r** | 0.226198 | -0.01057 | 0.210632 | 1 | 0.201659 |
|  | *p*-Value | 0.160467 | 0.948377 | 0.192027 |  | 0.212094 |
| **Zn** | **r** | 0.064467 | -0.27933 | 0.157212 | 0.201659 | 1 |
|  | *p*-Value | 0.692689 | 0.080885 | 0.332639 | 0.212094 | - |

## Discussion

Minerals and trace elements are essential requirements for physiological reactions in body metabolic processes [1]. Hence, alterations in their levels can have disastrous consequences in the body. Our study findings revealed that Type 2 Diabetes Mellitus (T2DM) exhibited a significant increase in the levels of Iron and Copper compared to healthy controls. Conversely, lower levels of Calcium, Magnesium, and Zinc were observed in T2DM patients compared to controls. These findings are consistent with previous studies [9-11]. However, several studies have demonstrated a lack of significant variation in serum zinc and magnesium levels between patients with T2DM and non-diabetic individuals, contradicting the results of the present study [12-13].

Zinc is a trace element that plays a vital role as a cofactor in over 300 metalloenzymes. Our findings revealed a significant reduction in zinc levels, consistent with studies by [4]. The process by which patients with T2DM become zinc deficient remains elusive and not fully understood. However, this could be due to the osmotic effect of glycosuria as the cause of urinary loss of this element.

Similar to previous research [14-17] which showed a significant difference in serum manganese levels, our study found a significant reduction between patients with T2DM and the control group. Although, a study was inconsistent with ours [11], it reported an insignificant difference between the two groups. Manganese is a vital component of certain enzymes, including hexokinase, enolase, and glycosyl transferases. Its proper level is required for insulin production and release.

Most studies conducted on serum calcium levels in both diabetic and non-diabetic subjects are insufficient. However, animal study of diabetes and studies of patients with diabetes indicate elevated levels of intracellular calcium in the majority of tissues[18]. Nevertheless, our study found a significant decrease in the diabetes group, which does not align with the work of [4] that reported no notable disparity in serum calcium among both T2DM and nondiabetic participants. The calciuresis can be attributed to hyperglycemia, which increases the nephron osmolality, thereby causing urinary loss of solute.

Studies have shown that higher levels of iron and ferritin in the blood are linked to insulin resistance[20]. Our study’s findings regarding serum iron concentration revealed a significant increase in iron levels, which were consistent with some previous studies [19, 21]. Excess iron levels lead to the generation of free radicals, resulting in oxidative damage to pancreatic beta cells. However, if not properly regulated and present in excessive amounts in the tissue, it can also lead to oxidative damage. Cells commonly contain the protein ferritin, which serves as an indicator of the body’s iron storage. The anomalous discrepancy in blood iron and ferritin levels in individuals with diabetes is enigmatic. However, insufficient consumption of food, lack of symptoms of the illness, and low rates of absorption are frequently linked to the reduction of iron and ferritin levels.

Previous studies have demonstrated an increase in serum copper levels among patients with T2DM [14,22,23,24], accompanied by high urinary copper excretion. Similarly, our study has revealed a substantial increase in the concentration of copper in the blood serum of patients with T2DM compared to those without diabetes. Copper is recognised as a potent catalyst for enzymes, and a lack of it leads to glucose intolerance, hypercholesterolemia, and atherosclerosis.

## Conclusion

Overall, our study observed changes in mineral concentrations, which highlight the importance of these micronutrients in maintaining metabolic balance and the potential consequences of their imbalance in T2DM. The results of our study are consistent with prior research, which indicates that individuals with type 2 diabetes mellitus (T2DM) have higher concentrations of iron and copper, while experiencing lower levels of calcium, magnesium, and zinc, in comparison to healthy individuals. These inconsistencies emphasise the importance of thorough monitoring of serum mineral levels in individuals with diabetes to gain a better understanding of disease progression and guide treatment approaches. The decreased concentrations of zinc and manganese in patients with T2DM suggest possible mechanisms related to urinary excretion and impaired enzymatic processes that are essential for insulin production and release. Conversely, the elevated blood levels of copper could potentially exacerbate metabolic issues and cardiovascular complications associated with diabetes. Furthermore, the observed differences in serum calcium and iron levels underscore the complex nature of mineral metabolism in diabetes. This emphasises the need for more research to understand the underlying mechanisms and the potential clinical consequences.

Ultimately, our research highlights the significance of dealing with mineral imbalances in the management of Type 2 Diabetes Mellitus (T2DM) and emphasises the necessity of specific interventions that aim to restore metabolic stability.

## Data Availability

All data produced in the present study are available on requests to the authors.

## Acknowledgment

Special thanks to Mohammed Alawami, the President of the ReachSci society for providing this invaluable opportunity for research training and development. We also acknowledge Khadijat Adefaye for her expert guidance in drafting this manuscript.

